# Black Women and the Preemie Prep for Parents (P3) Program: Exploratory Analysis of a Clinical Trial

**DOI:** 10.1101/2024.08.28.24312637

**Authors:** Siobhan M McDonnell, Kathryn E Flynn, Kris Barnekow, U. Olivia Kim, Ruta Brazauskas, S. Iqbal Ahamed, Jennifer J McIntosh, Michael B Pitt, Steven R Leuthner, Abbey Kruper, Mir A Basir

## Abstract

**Background:** The smartphone Preemie Prep for Parents (P3) program was developed to address the gap in prenatal education of preterm birth in high-risk pregnancies. Despite a higher incidence of preterm birth, Black women are less likely to receive prenatal education.

**Methods:** Pregnant women with medical conditions that predisposed them to preterm birth were randomized to receive the P3 program or links to American College of Obstetricians and Gynecologists webpages (control). The P3 group received periodic text messages, starting as early as 18 weeks gestational age, each with a link to a short, animated educational video. Participants completed the Parent Prematurity Knowledge Questionnaire, PROMIS Anxiety scale, and a feedback survey. This is a subgroup analysis of the Black, non-Hispanic participants in the P3 trial.

**Results:** Of the 26 Black non-Hispanic women enrolled, the P3 group (n=14) had higher knowledge scores than the control group (n=12), 67.5% correct vs. 43.6% (difference 24.0; 95% CI, 7.4 to 40.6), without experiencing an increase in anxiety. More P3 participants reported discussing preterm birth with their partner (100%) than control participants (57%; difference 43; 95% CI, 6 to 80).

**Conclusions:** The P3 program appears to be an effective method of providing preterm birth education to Black pregnant women.

## 1. Introduction

Preterm birth is the leading cause of US infant mortality and childhood morbidity.^1^ Preterm birth education is typically not provided to pregnant women until the delivery hospitalization.^2^ Waiting until the end of the pregnancy to provide education misses the opportunity to empower patients to make truly informed healthcare decisions.^3^ To address this gap, we developed the smartphone Preemie Prep for Parents (P3) program and tested it in a randomized controlled trial (RCT), showing greater preterm birth knowledge and preparedness to make key medical decisions throughout pregnancy.^4^ Despite a 50% higher incidence of preterm birth,^5^ Black women are less likely than other US women to receive prenatal education.^6^ While certain mobile health (mHealth) interventions have been explored in Black pregnant women, none directly address the preterm birth risks.^7-9^ Herein we examine the effect of the P3 program on preterm birth knowledge and preparation among Black pregnant women to make preterm birth healthcare choices.

## 2. Methods

This is an exploratory post-hoc subgroup analysis of a RCT (NCT04093492) approved by an institutional review board. Pregnant women with medical conditions that predisposed them to preterm birth (history of preeclampsia, chronic hypertension, diabetes requiring medications, intrauterine growth restriction, short cervix, multifetal gestation, and history of preterm birth) were recruited by phone from a high-risk obstetric clinic and randomized 1:1 to P3 or the control group. The randomization sequence was generated in R and implemented through Research Electronic Data Capture (REDCap).^10^ The P3 group received periodic text messages, starting as early as 18 weeks gestational age (GA) and continuing until preterm birth or 34 weeks GA. Each P3 text message had a link to 1 of 51 short, animated educational videos. The control group received links at the time of enrollment to American College of Obstetricians and Gynecologists patient education webpages. Study assessments at 25-, 30-, and 34-weeks GA included the Parent Prematurity Knowledge Questionnaire (PPKQ),^4^ a measure of core knowledge needed to participate in decisions about preterm birth. The PPKQ was refined through multiple rounds of cognitive interviews featuring diverse patient samples,^11, 12^ and is scored out of 100 percentage points. At each follow up, the Preparation for Decision Making Scale^13^ was administered, a 10-item measure of how well a decision aid prepared someone to communicate with clinicians about a health care decision. The scale is customized to a specific decision; we assessed the decision for neonatal resuscitation at 25 weeks GA, a risk appropriate birth hospital at 30 weeks GA, and breastfeeding choice at 34 weeks GA. The scale has been shown to discriminate between patients who did and patients who did not find a decision aid helpful, with a difference of 12 points on a 100-point scale and SD of 20.^13^ To explore experiential differences and potential harms of the education, the PROMIS Anxiety computerized adaptive test^14^ was collected at enrollment and each follow up assessment. The PROMIS Anxiety scale measures general anxiety over the past 7 days, with a population mean of 50 ± 10.

This post-hoc subgroup analysis examined responses from participants self-reporting as Black and not Hispanic or Latino. Hispanic or Latino participants were excluded from this analysis because of long-standing concerns of Hispanics being categorized by race,^15^ including many Hispanics viewing the ethnic identity as their racial identity^16^ and the demographic differences.^17^ Comparisons used t-tests for continuous variables and chi-square tests for categorical variables.

## 3. Results

Of the 122 Black non-Hispanic women contacted by phone, 61 responded and 26 (21%) enrolled (Table 1); this compares to a 35% enrollment rate in the overall sample.^4^ The Black women enrolled in the P3 group (n=14) had significantly higher PPKQ scores than the Black women in the control group (n=12) at 25 weeks GA (67.5% vs. 43.6%; difference 24.0 percentage points; 95% CI, 7.4 to 40.6 percentage points) and 30 weeks GA (76.5% vs. 52.9%; difference 23.5 percentage points; 95% CI, 6.9 to 40.1 percentage points), though this difference did not reach significance at 34 weeks GA (76.4% vs. 54.3%; difference 22.1 percentage points; 95% CI, -5.5 to 49.7 percentage points), Figure 1.

**Table 1.** Demographics of Black participants.

| **Participant characteristics, No. (%)** | **Total**<br>**n=26** | **P3**<br>**n=14** | **Control**<br>**n=12** |
| --- | --- | --- | --- |
| **Race** |  |  |  |
| Identified race as Black only | 23 (88) | 11 (80) | 12 (100) |
| Identified race as Black and white<sup>a</sup> | 3 (12) | 3 (20) | 0 |
| **Education** |  |  |  |
| ≤ High school diploma | 12 (46) | 5 (36) | 7 (58) |
| Some college | 10 (38) | 6 (43) | 4 (33) |
| 4-year degree | 1 (4) | 0 | 1 (8) |
| Graduate degree | 3 (12) | 3 (21) | 0 |
| **Marital status** |  |  |  |
| Married | 5 (19) | 5 (36) | 0 |
| Not married | 21 (81) | 9 (64) | 12 (100) |
| **Living situation** |  |  |  |
| Living with partner | 17 (65) | 10 (71) | 7 (58) |
| Not living with partner | 9 (35) | 4 (29) | 5 (42) |
| **Preterm birth risk factor<sup>b</sup>** |  |  |  |
| Pregestational diabetes | 3 (12) | 1 (7) | 2 (17) |
| Chronic hypertension | 10 (38) | 2 (14) | 8 (67) |
| History of preeclampsia | 2 (8) | 1 (7) | 1 (8) |
| History of preterm birth | 13 (50) | 9 (64) | 4 (33) |
| Short cervix | 1 (4) | 0 | 1 (8) |
| Multifetal gestation | 5 (19) | 3 (21) | 2 (17) |

<sup>a</sup> Participants could self-report more than one race from National Institutes of Health prespecified racial categories; in our sample, the only other race identified by Black participants was white.
<sup>b</sup> Preterm birth risk factor percentages may not add up to 100% as patients could have more than one risk factor.

**Figure 1.**
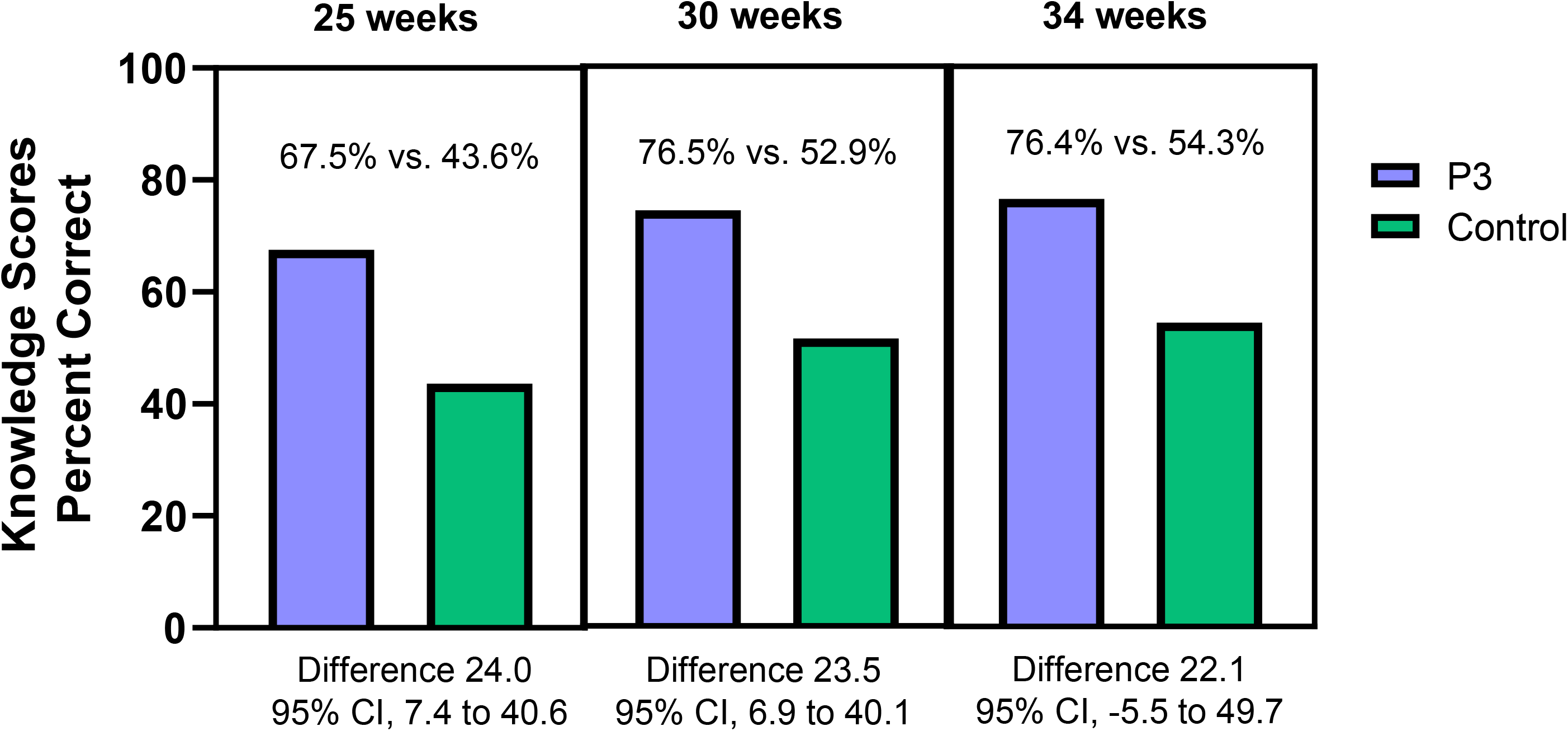
Knowledge scores of Black participants by study group.

Preparation for Decision Making Scale scores were higher in the P3 group than the control group at 30 weeks for a birth hospital choice (89.1 vs. 65.0, difference 24.1; 95% CI, 4.0 to 44.1). However, the difference did not reach significance at 25 weeks for a neonatal resuscitation decision (81.9 vs. 64.2; difference 17.7; 95% CI, -10.1 to 45.6) and 34 weeks for a breastfeeding decision (74.6 vs. 58.5; difference 16.1; 95% CI, -18.8 to 51.1), Figure 2.

**Figure 2.**
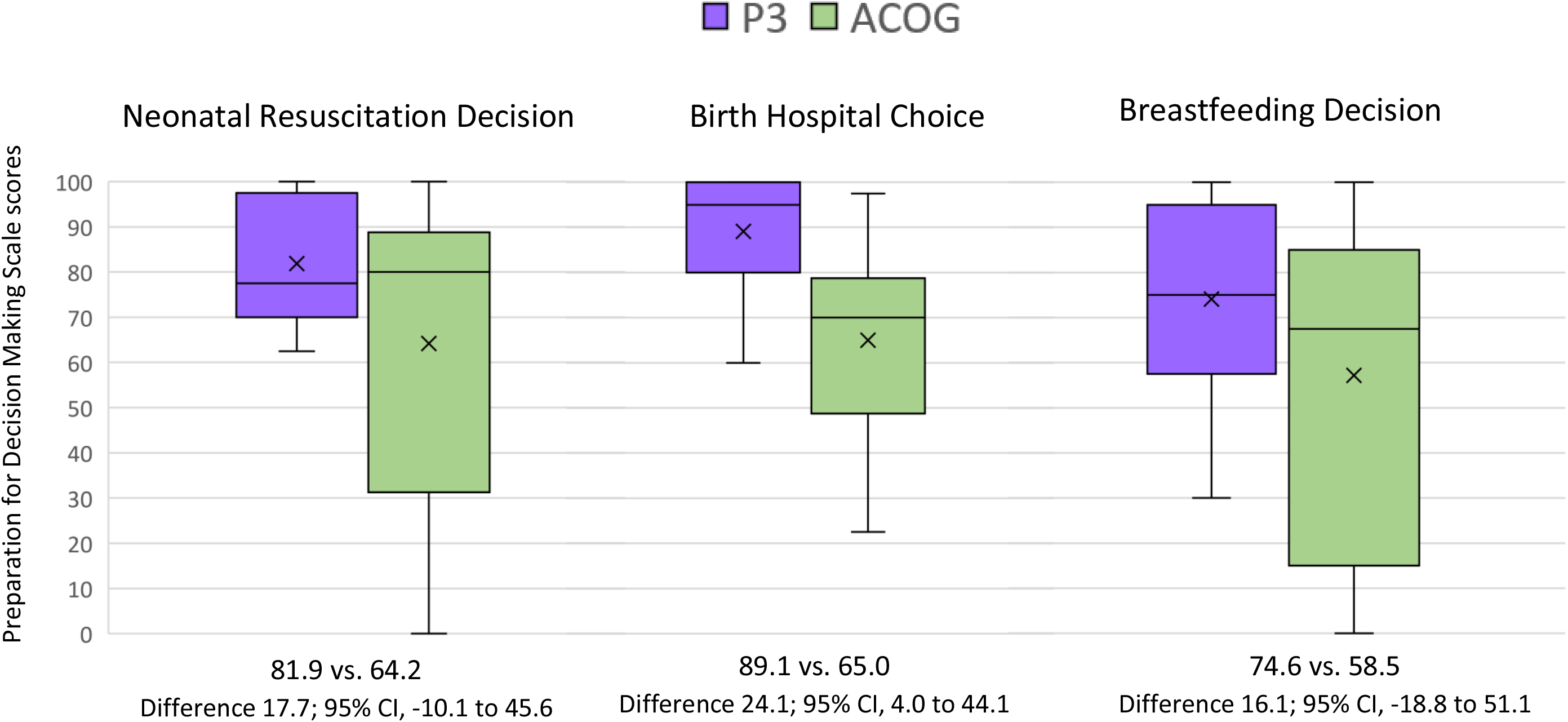
Preparation for Decision Making Scale scores of Black participants by study group.

Anxiety was not higher in the Black women in the P3 group than in the control group at enrollment (mean 56.1 vs. 52.8; difference 3.3; 95% CI, -4.2 to10.8), 25 weeks (52.9 vs. 50.6; difference 2.3; 95% CI, -7.2 to 11.8), 30 weeks (50.7 vs. 55.9; difference -5.2; 95% CI, -12.6 to 2.3), or 34 weeks (56.2 vs. 57.9; difference -1.7; 95% CI, -10.5 to 7.3) GA, Figure 3. The average Black P3 participant watched 30% of the 51 videos, compared to the overall P3 group mean of 55% of the videos watched.^4^

**Figure 3.**
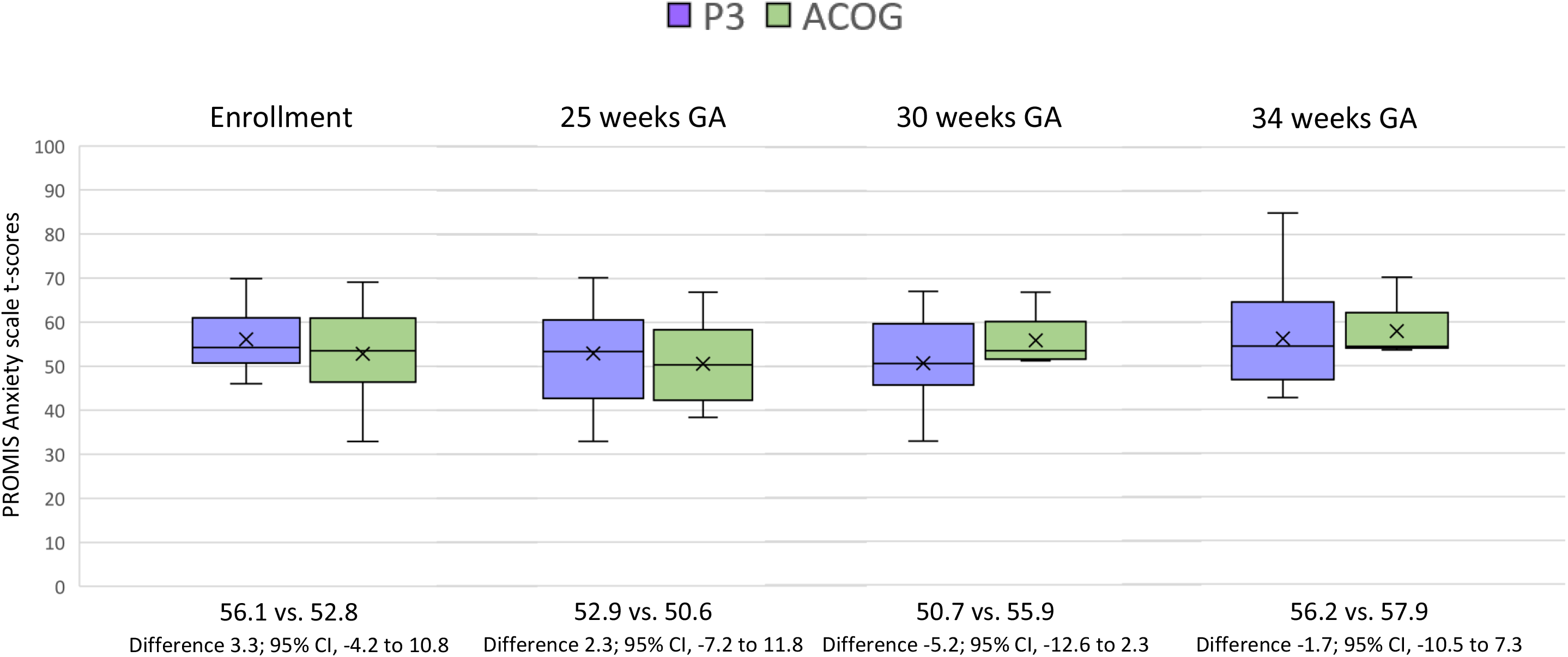
PROMIS Anxiety scale t-scores of Black participants by study group.

At study completion, more P3 participants reported discussing preterm birth with their partner (100% P3 vs. 57% control; difference 43 percentage points; 95% CI, 6 to 80 percentage points). Though not significantly different than the control group, more participants in the P3 group reported sharing preterm birth information with other family and friends (55% vs. 43%; difference 12 percentage points; 95% CI, -35 to 59 percentage points).

Of the P3 participants, 100% reported the P3 education provided more preterm birth information than their physician.

## 4. Discussion and Conclusion

### 4.1 Discussion

In this subgroup analysis of the Black women who participated in a RCT of the P3 program, those in the P3 group had better knowledge of preterm birth than those in the control group, without experiencing increased anxiety. Despite a limited sample, pregnant women in the P3 group were more prepared to make decisions regarding giving birth at a risk-appropriate birth hospital. Black women in the P3 group also reported having more discussions about preterm birth issues with their partners than those in the control group, and they all reported the program gave more preterm birth information than their doctor.

The racial disparity in preterm birth rates is largely due to downstream effects of structural racism.^18^ Black women face more barriers to clinic-based prenatal education^19^ because of issues such as transportation and childcare.^20, 21^ With further study, the P3 program may provide an option of circumnavigating these barriers to deliver preventive preterm birth education to those at higher risk of delivering preterm.^5^

This study was limited by its post-hoc analysis of a small sample and the low participation rate often seen among Black patients in clinical trials.^22^ While the trial’s sample matched the clinic’s demographics, the lower rates of engagement merit further exploration. Future studies to investigate the barriers and facilitators of P3 program usage will ensure greater acceptability and better tailor the program to Black families’ needs, as well as determine clinical outcomes.

### 4.2 Innovation

While smartphone-based prenatal education is widely used,^23^ providing preterm birth information through this modality is a new concept. This is reflected in the title given by Dr. Waldemar Carlo, a senior physician-scientist of Neonatal Research Network, in his *JAMA Pediatrics* editorial about the P3 program: “Preterm Prenatal Education—A Novel Approach.” ^24^ The P3 program offers a unique approach to prepare patients with high-risk pregnancies with information needed to improve outcomes. By offering prenatal preterm birth education to Black women through our smartphone-based platform, the P3 program overcomes transportation, childcare, and time off work-related barriers of traditional education for a group of patients particularly burdened by the clinic-based model. We are not the first to explore these potential benefits of mHealth for Black pregnancies.^7-9^ However, unlike the programs that include only normal pregnancies, the P3 program focuses on high-risk pregnancies and is the first mHealth intervention to provide education on preterm birth.

### 4.3 Conclusions

Even with suboptimal rates of engagement, the P3 program appears to be an effective method of providing preterm birth education to Black pregnant women. With further study, the inexpensive and easily scalable program could potentially benefit thousands of Black families in the US every year.

## Data Availability

Our research team will consider requests for re-use of scientific data from scientists of like-minded interests, and these will be prepared through MCW. No data sharing will take place until a signed data sharing agreement is in place, necessary costs of sharing provided, and all site PIs are aware and approve.

## Abbreviations

(NICHD): Eunice Kennedy Shriver National Institute of Child Health and Human Development
(GA): gestational age
(P3): Preemie Prep for Parents
(RCT): randomized controlled trial
(PROMIS): Patient-Reported Outcomes Measurement Information System
(REDCap): Research Electronic Data Capture

